# High-Throughput Digitization of Analog Human Echocardiography Data

**DOI:** 10.1101/2020.06.07.20123521

**Authors:** Alan C. Kwan, Gerran Salto, Emmanuella Demosthenes, Birgitta T. Lehman, Ewa Osypiuk, Plamen Stantchev, Ramachandran S. Vasan, Susan Cheng

## Abstract

Echocardiographic imaging data has been acquired for adult participants of a longitudinal community-based epidemiological cohort study at serial examinations between 1987 and 1998. The original image acquisition approach was analog with storage of moving images on Video Home System (VHS) tapes. Accordingly, we designed and implemented a standardized methodology for digitizing, anonymizing, and organizing these analog data to enable contemporary image-based analyses. Herein, we describe the overall methodology and the operational workflow, quality control, Health Insurance Portability and Accountability Act compliance, and data formatting issues that we addressed. We present this method as an accessible pipeline for enabling digitization of historical imaging data, originally acquired from large cohort studies, in order to preserve and repurpose them for application of advanced and continually evolving image analytical techniques. Such pipelines are critical not only for data conservation but are also invaluable for prospective analyses of imaging phenotypes in relation to already-accrued longitudinal outcomes in community-based cohorts that have been under careful surveillance over up to several decades of follow up.

## BACKGROUND

The longstanding Framingham Heart Study (FHS) has provided fundamental insights into the epidemiology of risk factors and cardiovascular disease – and a substantial portion of this seminal work has arisen from echocardiography.^1–3^ The Framingham Offspring Study was initiated in 1971 as a longitudinal community based cohort whose participants included the offspring of the original FHS cohort and their spouses.^4^ As part of the Offspring study, echocardiography was performed and recorded at multiple serial visits, including three visits that occurred between 1987-1998. At the time of original image acquisition, digital imaging was not broadly available. These initial imaging studies were recorded on Video Home System (VHS) tapes. It is known that VHS tapes tend to undergo up to 10-20% structural deterioration over a period of 10-25 years, depending on storage conditions.^5^ The echocardiographic information contained in these studies is a legacy of the FHS participants and their research efforts. It, therefore, warrants not only conservation but also facilitated accessibility for application of contemporary methods of analysis that offer the potential to derive new information from historical data. To this end, we worked to establish a process for converting echocardiographic videos captured on VHS tape into digital cine imaging files. We anticipate this endeavor will ultimately enable access of the data to multiple users and substantially reduce the physical storage space requirements of VHS tapes.^6^ Herein, we present our analog-to-digital conversion methodology, which we hope is generalizable as well as scalable to other cohort studies with historical echocardiographic data available. We recognize that application to other longstanding cohorts is of substantial interest given that historically collected imaging data can be related to prospectively already accrued outcomes data.

## METHODS

A team of core laboratory investigators developed and designed our analog-to-digital imaging data conversion process following a careful review and evaluation of previously reported methods. The finalized protocol was then approved by the FHS Executive Committee and our institutional review boards. As shown in **Figure 1**, the protocol involves a multi-step workflow. All VHS tapes are manually reviewed on a VHS tape player, with viewed content cross-checked against original logbooks and then physically labeled to verify which participant exams are recorded on a given tape. This workflow step allows for identification and correction of annotation errors that may have occurred during the initial recording and also optimizes an organizational process for locating studies. Each VHS tape contains a total of 8 to 24 studies, each of which varies slightly in duration as well as image quality (longer duration studies tend to include more recording time in order to accommodate for limitations of image quality). Following verified identification of a given participant study, we use a standard VCR machine connected to a specialized computer with commercial software optimized for facilitating analog-to-digital conversion of the VHS moving images to a digital format (TIMS 2000 medical computer, Foresight Imaging, Chelmsford, MA, USA). We select the Digital Imaging and COmmunications in Medicine (DICOM) format given its wide acceptance as a universal standard file format for medical imaging and the ability to share DICOM image files within a Picture Archiving and Communications System (PACS).^7^ We manually identify the highest-quality 4 to 6 cardiac cycles of each standard echocardiographic view during the digitization process, with each clip recorded digitally as separate recording unit and saved as a separate DICOM file within the corresponding study. We make note of those views deemed to have inadequate quality, in a corresponding digitization logbook (i.e., laboratory notebook), although we retain the total number of 4 to 6 cardiac cycles selected and recorded to maintain study record consistency. Finally, for a given participant examination, we manually upload the newly generated DICOM files from the local hard drive on the TIMS system to our PACS residing within a centralized and firewall secured networked server environment. Overall, a total of approximately 10 to 20 cine clips are recorded per study, depending on the availability and quality of images at hand.

**Figure 1.**
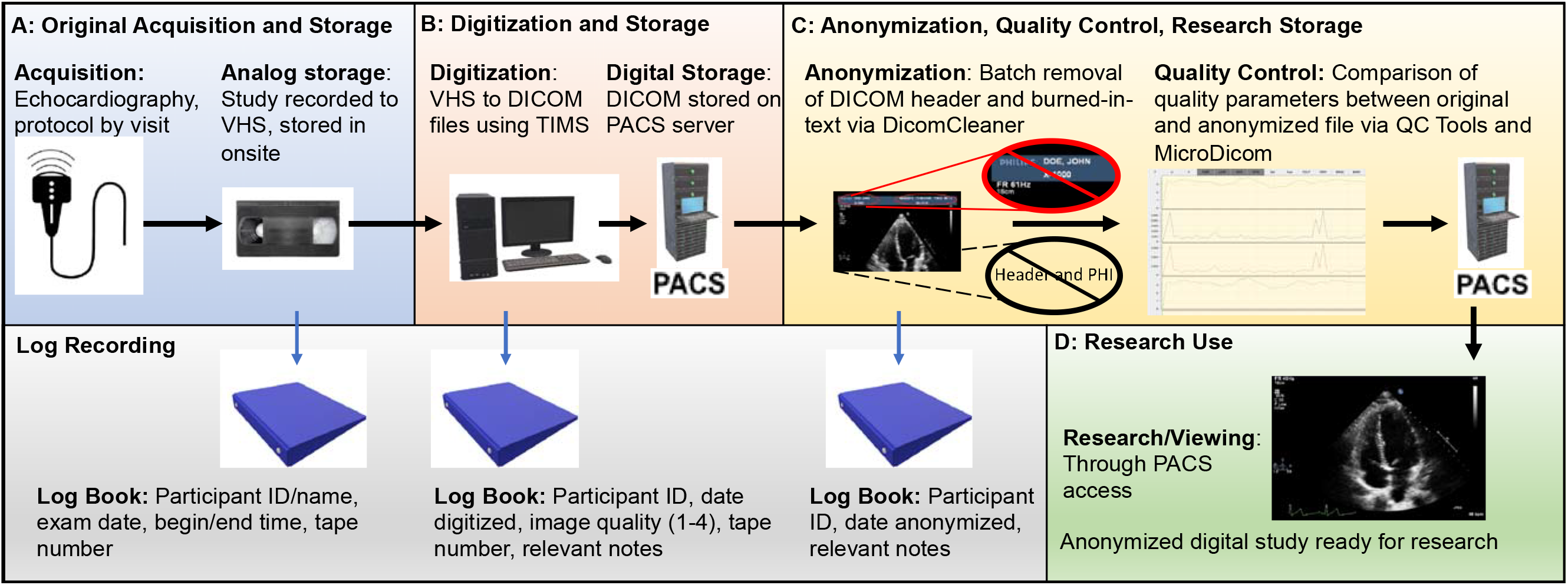
Methods diagram from original acquisition and storage of image, digitization, anonymization, and research pipeline. VHS: Video Home System, DICOM: Digital Imaging and Communications format in Medicine, PACS: Picture Archiving and Communications System.

### Framingham Offspring Cohort

The Framingham Offspring cohort study is one of the nation’s earliest second-generation epidemiologic studies.^8^ The Offspring cohort study began in 1971, as an expansion from the original 1948 Framingham Heart Study, by enrolling the original cohort’s offspring as well as their spouses.^9^ Participants underwent regular examinations on average every four years for the duration of the participant’s life. A total of 5,124 participants were originally enrolled in this study, with currently 2,675 living participants still enrolled.

### Original Image Acquisition and Storage

Echocardiographic imaging, using analog video recording methods, was included as part of the Framingham Offspring cohort examinations that were the focus of the current study beginning in 1987 (**Table 1**), prior to wider acceptance of standardized digital storage formats (**Figure 2**).^10^ For this reason, analog recording to VHS was used not only for the initial exams but also continued through several subsequent cycles of exams for the same participants.^11^ The echocardiographic image acquisition protocol was standardized for each examination of the Offspring cohort examinations 4, 5 and 6 using an HP SONOS 1000 ultrasound machine equipped with a Panasonic AG6300 VHS video recorder (**Table 2**). The VHS tapes were stored onsite in a secured, temperature and humidity-controlled facility. Corresponding logbooks containing participant name, cohort study identification number, exam number, tape number, date of exam, and exam beginning and ending time were maintained.

**Figure 2.**
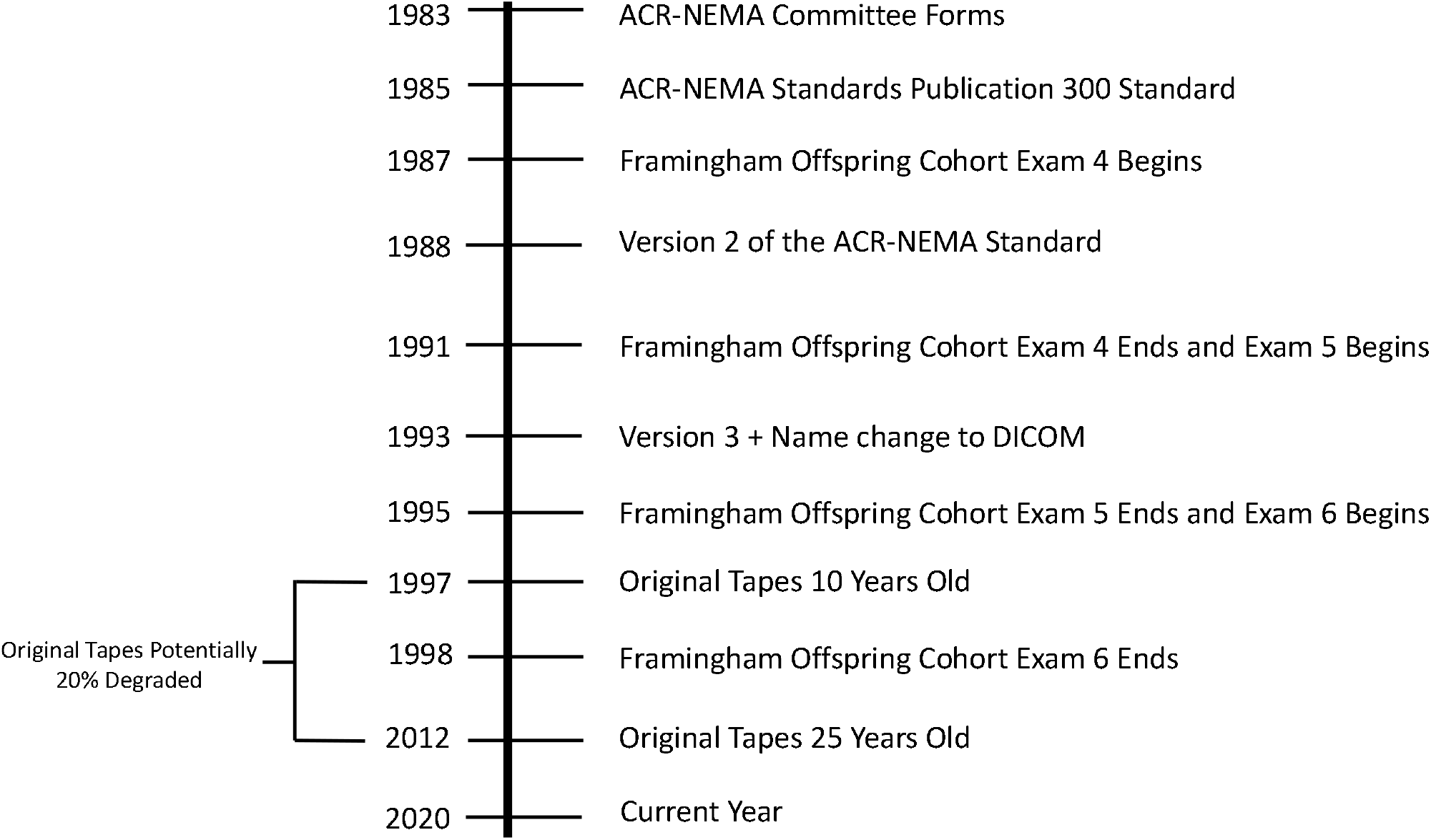
Timeline of Framingham Offspring Cohort and digital storage standards. In 1983 ACR (American College of Radiology) and NEMA (National Electrical Manufacturers Association) form ACR-NEMA Committee to create an imaging standard that satisfied the needs of both physicians and medical imaging equipment manufacturers. Three versions of the standard were released. The first in 1985, the second in 1988, and the third in 1993.

**Table 1.** Dates of original examinations, exam conversion, details, and considerations

|  | Original Study<br>Recording Dates | # Cardiac Cycles | Variable Considerations |
| --- | --- | --- | --- |
| Exam 4 | 1987-1991 | • 4 to 6 cardiac cycles per 2D view<br>• 3 cardiac cycles per m-mode, PW, and CW image | • Quality of study - affected # of cycles obtained and images obtained<br>• Length of study - affected # of cycles obtained<br>• Availability of images - affected which images we could obtain |
| Exam 5 | 1991-1995 |  |  |
| Exam 6 | 1995-1998 |  |  |

**Table 2.**
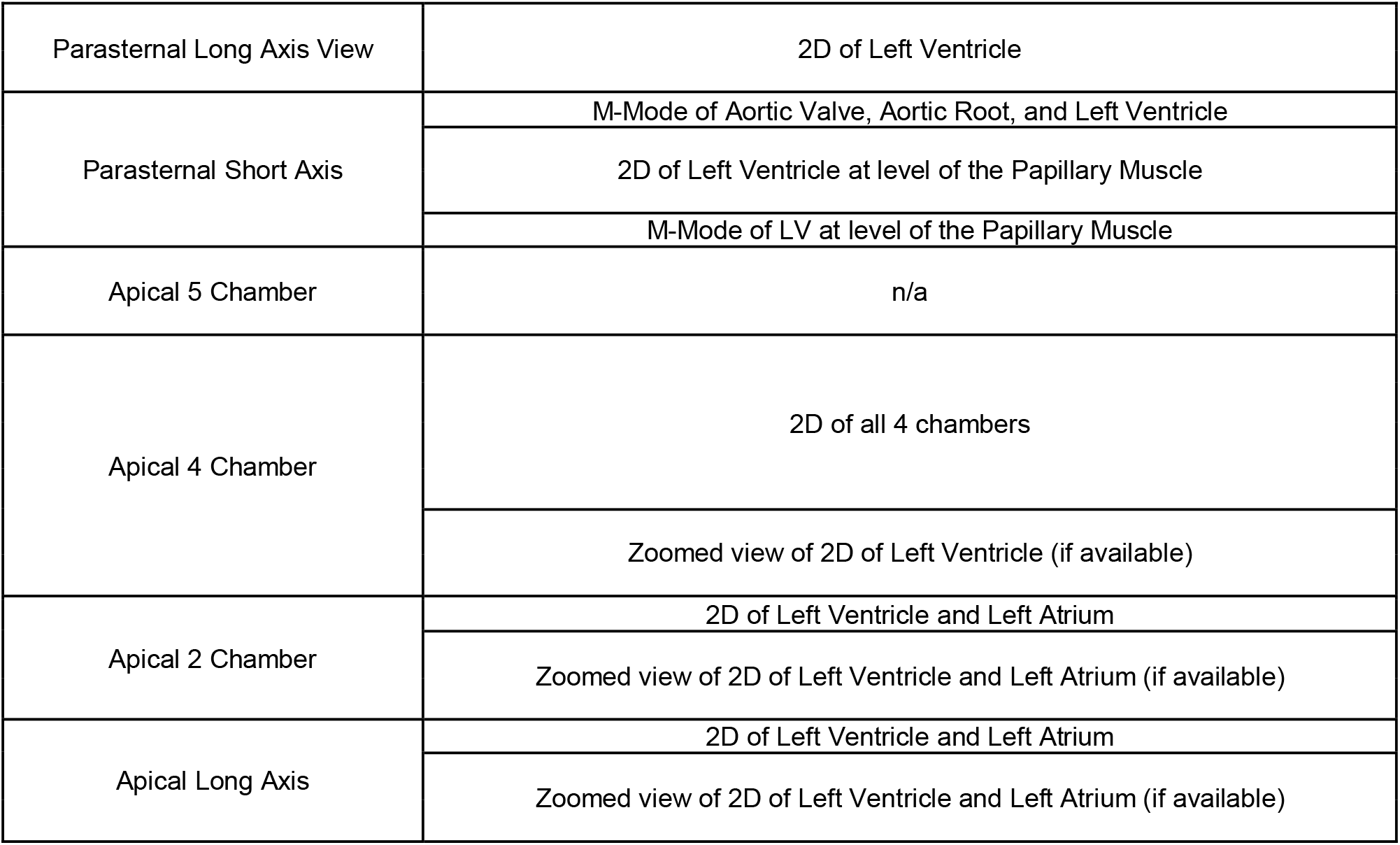
General Digitization Protocol. LV = left ventricle, RV = right ventricle, TV = tricuspid valve, MV = mitral valve, TR = tricuspid regurgitation, AR = aortic regurgitation, MR = mitral regurgitation, LVOT = left ventricular outflow tract CW = continuous wave, PW = pulse wave

|  |  |
| --- | --- |
| Parasternal Long Axis View | 2D of Left Ventricle |
| Parasternal Short Axis | M-Mode of Aortic Valve, Aortic Root, and Left Ventricle |
|  | 2D of Left Ventricle at level of the Papillary Muscle |
|  | M-Mode of LV at level of the Papillary Muscle |
| Apical 5 Chamber | n/a |
| Apical 4 Chamber | 2D of all 4 chambers |
|  | Zoomed view of 2D of Left Ventricle (if available) |
| Apical 2 Chamber | 2D of Left Ventricle and Left Atrium |
|  | Zoomed view of 2D of Left Ventricle and Left Atrium (if available) |
| Apical Long Axis | 2D of Left Ventricle and Left Atrium |
|  | Zoomed view of 2D of Left Ventricle and Left Atrium (if available) |

### Storage Cross-Check and Digitization

On recovery from storage, VHS tapes are individually viewed and cross-checked against original logbooks. A Mitsubishi MD3000 VCR connected to a TIMS 2000 medical computer (Foresight Imaging, Chelmsford, MA, USA) system is used to convert VHS data to DICOM data. The DICOM header is populated during the TIMS conversion process with participant name, ID number, and study date.^12^ Images are captured separately under the relevant study and edited, as needed, to store the best quality 4-6 cardiac cycles of each view, excluding any static or artefacts. For each study that is full processed via the digitization workflow, the final digitized images are sent to our PACS system and stored on a local secured access server. Each study requires 15-20 minutes to digitize manually, depending on the quality and technical issues pertaining to each individual echocardiographic study. Corresponding logbooks containing participant ID, digitization date, digitization quality (graded on a scale of 1 to 4), tape number, and any relevant notes are maintained.

### Anonymization

Healthcare Insurance Portability and Accountability Act (HIPAA) mandates removal of protected health information from any medical records before use in a research context. DICOM files must be anonymized in the text-based header and by removal of any text within images. Proprietary software (coded in Python v.3.8.1) is used to anonymize images in batches of 20, whereby all 20 studies are stripped of headers at the same time. Then, 2D images and M-mode/doppler images are stripped of burned-in text separately, due to variation in the location of the text on the image itself. We remove unreplaced identities, descriptions, series description, acquisition protocol name, patient characteristics, all UID’s, unsafe private attributes, device identifiers, institution identifiers, clinical study attributes, all structured content, and unsafe structured content. Corresponding logbooks containing participant ID, anonymization date, and any relevant notes are maintained.

### Image Quality Control

Both the original DICOM file and the newly anonymized DICOM file are imported into MicroDicom and converted to AVI video files with no compression. Both AVI files are exported and stored in a designated folder for studies used for quality control. The AVI file of the anonymized image is labeled “[participant ID] – anon.” and the AVI file of the non-anonymized image is labeled “[participant ID] – original.” Both of these files are then imported into QC Tools where they are analyzed for major discrepancies using the metrics listed in **Table 3**. Quality control is performed on the first 10 studies and approximately once every 250 to 500 studies throughout the anonymization process.

**Table 3.** Image quality assessment variables. PSNR = Peak signal to noise ratio, MSE = Mean square error.

| IMAGE QUALITY ASSESSMENT |  |
| --- | --- |
| Metric | Information Given |
| Y-Value | Information about brightness and luminance |
| U-Value | Information about video color |
| V-Value | Information about video color |
| Y, U and V Diffs | Indicates the extent of visual change from one frame to the next |
| PSNR | Comparison as a ratio of the peak signals expressed as dB |
| Y-Range | Indicative of contrast range |
| Y, U, V averages | Average of Y, U and V values |
| MSE | Mean Square Error for each (YUV) plane. Higher values may be indicative of differences between 2 images being compared. |
| File Size | Differences in file size could be indicative of differences in video quality |

## RESULTS

Anonymization of the DICOM files is necessary prior to application of any new image analysis approach. Although a variety of DICOM anonymization software products are available online; performance of the different software packages vary widely.^13^ The anticipated total quantity of Framingham Heart Study studies is greater than would be feasible for manual deidentification. DicomCleaner (v1.0, PixelMed Publishing, Allentown, PA, USA) was chosen to deidentify DICOM file headers and redact burned-in text in batch quantities. Loss of image quality during anonymization is a concern due to the need to directly alter image data in order to remove patient identifiers within images. To ensure image quality, we compared anonymized and non-anonymized image quality. DICOM was converted to AVI using MicroDicom Viewer (MicroDicom, Sofia, Bulgaria) and compared image quality with QC Tools (V1, by MediaArea, Curienne, France). We compared the 2 video files for changes in color, brightness, contrast, luminance, and peak signal-to-noise ratio. On quantitative and qualitative review, we did not observe any significant differences between anonymized vs non-anonymized studies.

The study pipeline consists of multiple steps in order to ensure adequate quality, accurate labeling, and complete anonymization. While digitization itself has been straightforward, quality assurance with respect to image quality and complete anonymization comprise the most time-and effort-intensive steps. We recognize the essential priority of ensuring adequate subject protection and data quality. To this end, project pipeline work including quality assurance procedures are ongoing for the full compendium of available studies. Current and future plans are in place to perform quantitative analysis on the data including conventional and advanced image analysis using approaches such as texture analysis, machine learning, and deep learning. We anticipate that multiple projects will be feasible using these data. Data sharing will be enabled from a centralized secure server via standard institutional data sharing agreements.

## DISCUSSION

Conversion of archival analog echocardiography to research-ready digital storage is possible using a combination of proprietary and open source methods. Physical storage differences are substantial with associated cost savings, given the resources required for both space and maintenance of both analog media as well as analog reading devices. Digitally stored data allows for access by multiple users and locations and is easily duplicated for analyses. Current methods for ensuring digital fidelity and protection against data loss via hardware degradation include storage of servers and hard drives in temperature-controlled rooms, performance of regular maintenance on storage hardware, protection from electrical surges, and use of high quality equipment.^12,14,15^ All of these methods of protection ensure that the lifetime of the Framingham Offspring Study digital data will extend far beyond the initial storage methods for future research.

Cardiac disease remains the most common cause of death in the United States.^16^ While multiple modern cohorts collect digital cardiovascular imaging data, the use of data from historical cohorts have an advantage of previously accrued outcomes data that can be immediately analyzed. Analog quantitative echocardiography data from Framingham Offspring Cohort has been assessed using conventional as well as advanced image analysis methods, as reported in multiple prior publications.^17–20^ Digital formatting of images will allow for alternative methods of analysis including texture analysis and artificial intelligence approaches.^21^ Given the large quantity of clinical and outcomes accrued over time, the digitally-converted imaging data are likely to be appropriate for supervised deep learning analyses.

An improved understanding of cardiac disease remains critical for population health. Historical cohorts with completed enrollment have long-term outcomes data. There have been multiple historical cohorts that have collected analog echocardiography data, ranging in size up to over 5000 participants and dating back to 1988. Few studies have digitized their historical data. The HyperGEN Archeological Echocardiography study has had the largest digitization process to date. This study examined association between hypertension genetics and left ventricular hypertrophy, digitized 2150 of 2234 echoes and applied strain analysis.^22^ The HyperGEN study used a similar TIMS setup with offline storage and no formal anonymization process described. Other alternative approaches to digitize echocardiograms stored on analog tapes exist using alternative conversion hardware and electronic storage methods.^23, 24^ We offer our experience at the Framingham Heart Study as another proof of concept that preservation of legacy data can be cost-effective, time efficient, consistent, and scalable, in organization of data in a research-ready durable storage format.

We recognize limitations within our workflow. All processes were performed at a single site with the same equipment by a dedicated research team. Therefore, we are uncertain of the flexibility of this model to be generalized to alternative locations with differing infrastructure. The quantity of time and use of manual processes remains a limitation, as this required a full-time team of research staff and multiple years of effort. The manual process of logging and labeling is at risk for error, which could affect data labeling. At this time, we have completed digitization, quality assurance procedures, and anonymization for a proportion of the total planned number of studies and, thus, additional limitations may become apparent with further progress.

Large community-based epidemiological cohort studies with historically collected data continue to offer a multitude of opportunities to derive new insights relevant to modern medical questions. As the accessibility to and support for analog data decrease over time, a number of large cohort studies with historically collected analog imaging data will face similar challenges related to data conversion, anonymization, quality control, research preparation, and storage, for which we offer a scalable approach. Further automation of this process is being pursued. Preservation of the data, and storage in a format amenable to novel research methods is a worthy goal for investigators. Our method is scalable and maintains image quality across contemporary storage formats and may provide a blueprint for future efforts to maintain and update historical imaging data collected from clinical research studies.

## Data Availability

No data will be generated directly from this methods and protocol development project.

